# Patient attitude towards lecanemab: results from a survey within the European Alzheimer’s disease consortium (EADC), the German memory clinic network (DNG) and Austrian memory centers

**DOI:** 10.1101/2025.06.16.25329507

**Authors:** Jonathan Vöglein, Johannes Levin, Elisabeth Stögmann, Christian Haass, Günter U. Höglinger, Lutz Frölich, Frank Jessen

**Affiliations:** Department of Neurology, LMU University Hospital, LMU Munich, Marchioninistraße 15, 81377 Munich, Germany; German Center for Neurodegenerative Diseases (DZNE), Feodor-Lynen-Straße 17, 81377 Munich, Germany; Munich Cluster for Systems Neurology (SyNergy), Feodor-Lynen-Straße 17, 81377 Munich, Germany; Deutsches Netzwerk Gedächtnisambulanzen, Pauwelsstraße 30, 52074 Aachen, Germany; Department of Neurology, Medical University of Vienna, Währinger Gürtel 18-20, 1090 Vienna, Austria; Comprehensive Center for Clinical Neurosciences & Mental Health, Medical University of Vienna, Währinger Gürtel 18-20, 1090 Vienna, Austria; Biomedicine Center (BMC), Biochemistry, LMU Munich, Großhaderner Straße 9, 82152 Planegg, Germany; Department of Geriatric Psychiatry, Central Institute of Mental Health Mannheim, Medical Faculty Mannheim, University of Heidelberg, Heidelberg, J5, 68159 Mannheim, Germany; European Alzheimer’s disease Consortium, Kerpener Straße 62, 50931 Cologne, Germany; Department of Psychiatry, University of Cologne, Medical Faculty, Kerpener Straße 62, 50931 Cologne, Germany; German Center for Neurodegenerative Diseases (DZNE), Bonn/Cologne, Venusberg-Campus 1, 53127 Bonn, Germany

**Keywords:** Alzheimer’s disease, lecanemab, anti-amyloid antibodies, patient preferences, regulatory approval, APOE4 homozygosity, risk-benefit perception, patient survey, shared decision-making, disease-modifying therapy

## Abstract

**INTRODUCTION:** Lecanemab approval was long pending in the EU but has now been granted. European patient attitudes towards anti-amyloid therapies are insufficiently assessed and reported.

**METHODS:** We conducted an anonymous, multicenter survey across European memory clinics targeting patients with early symptomatic AD. A standardized questionnaire with four yes/no questions assessed attitudes toward lecanemab treatment and approval, including in the context of APOE4 homozygosity.

**RESULTS:** Among 281 participants, endorsement rates were high for both individual treatment (82%) and general EU approval (92%). While still high, support was lower for treatment and approval in the context of APOE4 homozygosity (61% and 77%). Support for approval for APOE4 homozygotes declined after regulatory recommendations excluded this group. Results were consistent across European regions.

**DISCUSSION:** Strong support for access to lecanemab reflects the significant patient need for disease-modifying therapies and underscores the importance of considering patient perspectives in regulatory decision-making.

## 1 Background

Lecanemab is a monoclonal antibody directed against amyloid-β that effectively clears amyloid-β plaques from the brain [1]. In a phase 3 trial, lecanemab slowed down clinical progression of Alzheimer’s disease (AD) by about 30% in 18 months [1]. As a result, lecanemab was approved for treatment of early symptomatic Alzheimer’s disease in several regions worldwide, including the USA [2], Japan [3], China [4], South Korea [5], Hong Kong [6], Israel [7], the United Arab Emirates [8] and the UK [9]. In the UK, however, approval was limited to non-carriers or heterozygotes of the apolipoprotein E (APOE) ε4 genotype, only.

Treatment with lecanemab can be associated with amyloid related imaging abnormalities (ARIA), a class-specific side effect of monoclonal amyloid-β antibodies. In the phase 3 trial of lecanemab, ARIA occurred in approximately 12% of treated patients [1]. In one quarter of these 12%, i.e. in about 3% of all patients treated with lecanemab, ARIA were accompanied by symptoms. Whereas no deaths associated with lecanemab occurred in the core phase 3 trial, in the open-label extension phase ARIA were associated with fatal outcomes in isolated cases [10]. In an opinion of 25 July 2024, the Committee for Medicinal Products for Human Use (CHMP) of the European Medicines Agency (EMA) considered the benefit/risk-ratio of lecanemab not sufficient and recommended against an approval of the drug in the European Union (EU) [11]. After a re-examination procedure, the CHMP adopted a positive opinion published on November 14, 2024. Similar to the UK, this positive opinion restricted eligibility to individuals who are not homozygous for APOE ε4 [11]. After a second re-examination considering newly available safety data, the CHMP concluded that its November opinion did not require revision. Consequently, the European Commission resumed the decision-making procedure regarding the market authorization of lecanemab [11]. Finally, EU approval was granted on April 15, 2025 [12].

The course of this evaluation process has sparked several position statements from various stakeholder groups. Scientists [13,14], scientific journals [15], medical/scientific societies [13,16,17], publicly funded research organizations [18], patient advocacy groups [19,20,21], the drug’s manufacturer [22], and the media [23,24] have mostly voiced concerns or expressed difficulty understanding CHMP’s initial negative opinion.

However, knowledge about the attitudes toward lecanemab of one key stakeholder group is missing: patients affected by early symptomatic AD who are potentially eligible for lecanemab treatment. To address this gap, we conducted a patient survey across Europe.

## 2 Methods

### 2.1 Participants

Patients with mild cognitive impairment or mild dementia and a confirmed positive amyloid-β status, that is, patients with early symptomatic AD potentially suitable for treatment with lecanemab, were eligible to participate in the survey. Eligibility was determined by treating neurologists and/or psychiatrists.

The survey was conducted anonymously and collected no personal or demographic data. The ethics committee of LMU Munich confirmed that no formal consultation was required for conducting this anonymous survey (reference number: 24-0894-KB).

### 2.2 Survey design

The primary goal for the survey design was to convey information about the benefits and risks of lecanemab as simply as possible, using lay language appropriate for cognitively impaired individuals, while ensuring sufficient detail to enable well-informed responses. The survey was designed by JV, FJ, and LF, with advisory input from JL. The design process involved both email correspondence and several in-person meetings. The survey was administered using the online platform SurveyMonkey. Figure 1 illustrates the final survey design and wording of the English Version. Completion of the survey required answering all four questions.

**Figure 1:**
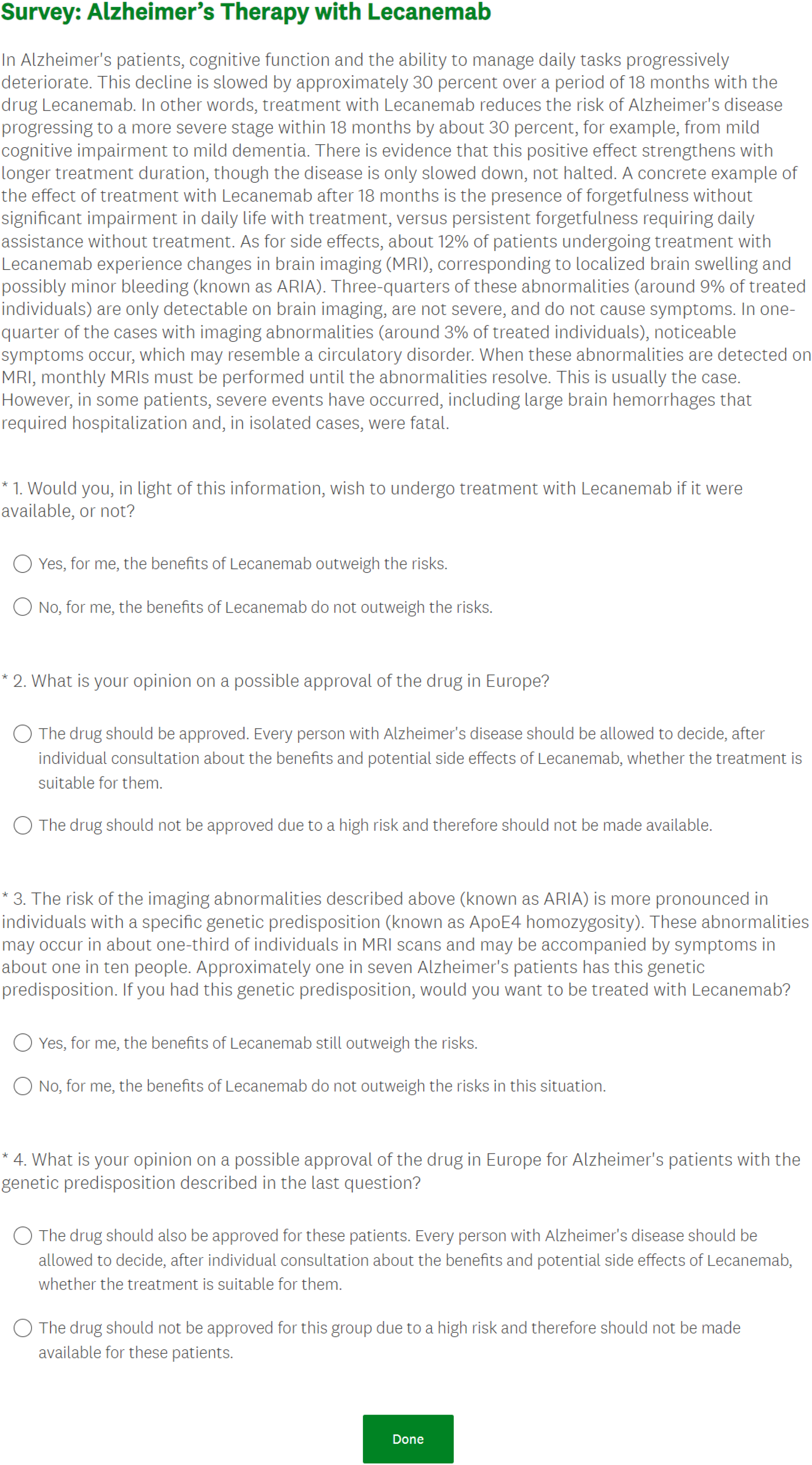
Survey appearance and wording in English.

### 2.3 Survey distribution

The survey was distributed through memory clinics of the German Network of Memory Clinics (Deutsches Netzwerk Gedächtnisambulanzen, DNG) [26], memory clinics in Austria affiliated with the departments of neurology at medical universities in Vienna, Graz, Innsbruck and Salzburg, and within the European Alzheimer’s Disease Consortium (EADC) [27]. The DNG and the EADC are non-profit medical and scientific organizations. The DNG provides a national network for AD centers in Germany [26], while the EADC promotes collaborative AD research across multiple European countries [27]. Standardized versions in German language were used for Germany and Austria, and a standardized English version for the EADC. After eligible patients were identified by their treating physicians, a survey link was provided. For patients lacking the necessary technical skills or resources to complete the online survey, a paper version could be filled out manually and later entered into the online system by the treating physician. For patients not fluent in English, EADC centers had the option to translate the English survey version into local languages using an artificial intelligence-based translation tool. The survey was launched on October 14, 2024, in the DNG, and on October 16, 2024, in Austria and the EADC. Common closing was on February 18, 2025.

### 2.4 Statistical analysis

For each of the four survey questions, binomial tests were conducted to assess whether the proportion of “yes” or “no” responses differed from chance (p = 0.5).

Fisher’s exact test was used to examine the influence of question type (treatment-vs. approval-related questions), population context (general vs. APOE44), target population (DNG, Austrian centers, EADC) and timepoint of survey completion (before vs. after CHMP’s positive opinion on November 14 2024) on responses. Fisher’s exact test was chosen over regression models because it is more appropriate for small or imbalanced subgroup sizes and handles complete separation (e.g., 100% endorsement for lecanemab approval for APOE44-carriers in Austria) without instability. All tests were two-sided. P-values < 0.05 were considered statistically significant. Statistical analyses were conducted using R version 4.3.3.

## 3 Results

### 3.1 Survey design and language

The appearance and wording of the survey are shown in Figure 1. In brief, after an introduction describing the benefits and risks of lecanemab considered relevant for decision-making, four yes/no questions addressing the following topics were asked: 1. the wish to be treated with lecanemab; 2. the opinion on the approval of lecanemab in the EU; 3. the wish to be treated with lecanemab in the hypothetical case of APOE4 homozygosity; 4. the opinion on the approval of lecanemab for APOE4 homozygotes in the EU.

### 3.2 Participants

A total of 281 patients with early symptomatic AD completed the questionnaire. The response rate was not assessed in this anonymous survey. EADC-affiliated sites contributed 202 patients (72%), 60 (21%) were from DNG centers, and 19 (7%) from memory clinics in Austria (Table). The EADC population included, but was not limited to, responses from Belgium, Bulgaria, Czech Republic, Greece, France, Italy, Portugal, Slovakia, Spain and Sweden.

**Table:**
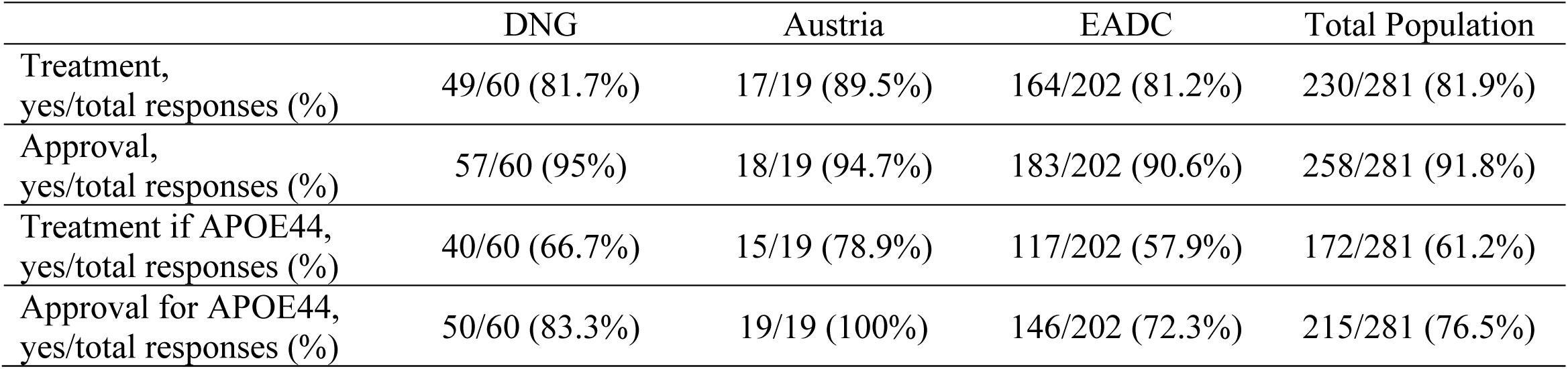
Population numbers and survey responses regarding lecanemab treatment and approval stratified by region. Abbreviations: DNG = Deutsches Netzwerk Gedächtnisambulanzen (German network of memory clinics); EADC = European Alzheimer’s Disease Consortium

### 3.3 Survey results

Endorsement rates were statistically significantly greater than expected by chance (i.e., 50%) for all four questions. A total of 81.9% of survey participants indicated that they would wish to receive lecanemab if it were available (230/281; 95% confidence interval (CI): 76.8–86.2%, p < 0.001). Support for EU approval of lecanemab was even higher, with 91.8% in favor (258/281; CI: 87.9–94.7%, p < 0.001). When asked about treatment preference in the case of known APOE4 homozygosity, 61.2% expressed a wish to be treated (172/281; CI: 55.2–66.9%, p < 0.001). Approval of lecanemab for APOE4 homozygous AD patients was endorsed by 76.5% of respondents (215/281; CI: 71.1–81.3%, p < 0.001) (Table, Figure 2).

**Figure 2:**
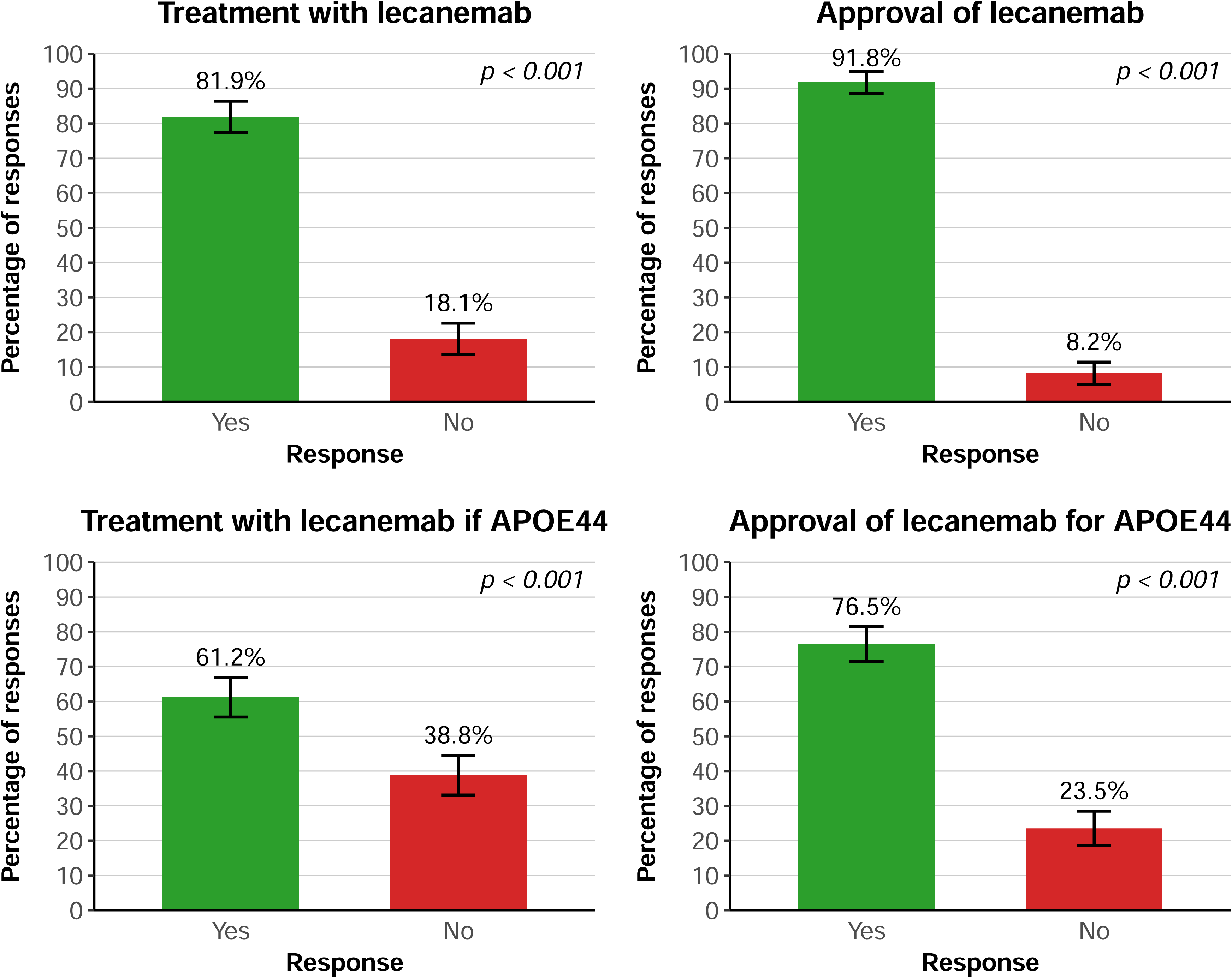
Survey responses regarding lecanemab treatment and regulatory approval. Description: Panel titles reflect the topics addressed in the corresponding survey questions. The majority of individuals with early symptomatic Alzheimer’s disease supported individual treatment with lecanemab and its approval in the European Union (EU). While endorsement remained high, it was lower for treatment willingness in case of APOE44 homozygosity and for EU approval for APOE44 carriers. Binomial tests showed that endorsement rates for all four questions were significantly above chance (50%), with p < 0.001 in each case. Error bars indicate 95% confidence intervals. Abbreviations: APOE44 = Homozygous carrier/carriers of the apolipoprotein 4 allele

Approval-related questions received higher endorsement than treatment-related ones, and questions framed in the general population context more than those targeting APOE44 carriers.

### 3.4 Response patterns

To explore how individual participants responded across all four survey questions, response combinations were analyzed. The most frequent pattern, endorsed by 56.6% of respondents, included affirmative answers to all four questions. Other response patterns were markedly less common. For example, 11.7% of participants supported approval for all early AD patients, including APOE4 homozygotes, and wished to be treated with lecanemab in the case they were APOE44 non-carriers, but not if they were hypothetically APOE44 homozygous. Complete non-endorsement across all four items was rare and observed in only 6.8% of participants.

Patterns with mixed responses confirmed the stronger endorsement of approval questions over treatment questions, and of general population items over those targeting APOE44 carrier questions. Figure 3 illustrates the response combinations observed in more than 5% of participants.

**Figure 3:**
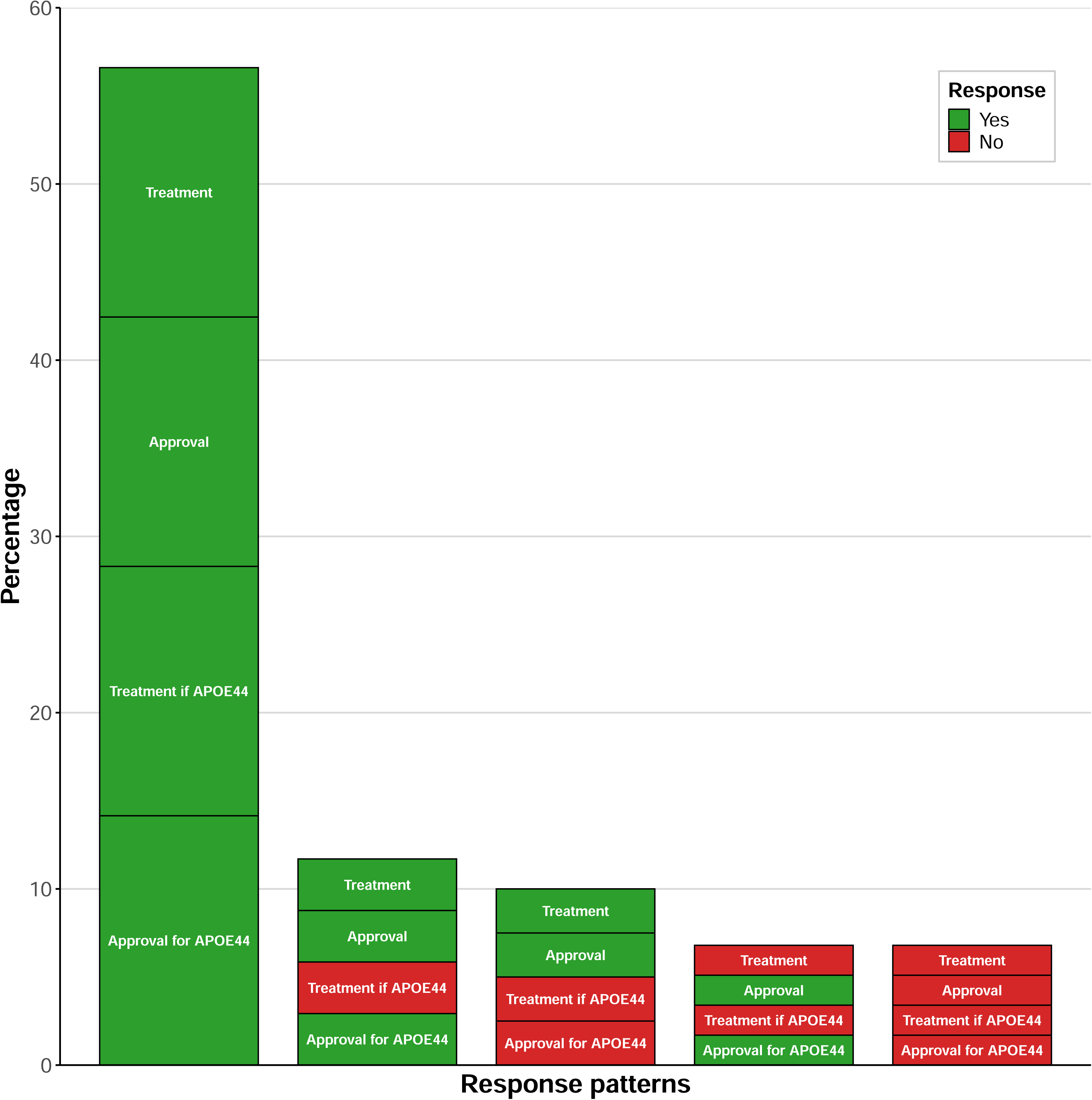
Survey response patterns. Percentages of the top five response combinations observed in the study are shown. The most frequent pattern by far (56.6% of participants) was full endorsement of both individual treatment and regulatory approval, in both general and APOE44-specific contexts. Abbreviations: APOE44 = Homozygous carrier/carriers of the apolipoprotein 4 allele

### 3.5 Survey outcomes by question type (treatment vs. approval)

To further investigate the observed trend of higher endorsement of approval-related compared to treatment-related questions, we grouped questions by targeted topic and conducted statistical testing. Endorsement for approval questions was statistically significantly more frequent (general context (irrespective of APOE status): 92%; APOE44 context: 77%) than for hose addressing treatment (general context: 82%; APOE44 context: 61%) (odds ratio [OR] = 2.48, 95% CI [1.44, 4.40], p < 0.001) (Figure 4, left panel).

**Figure 4:**
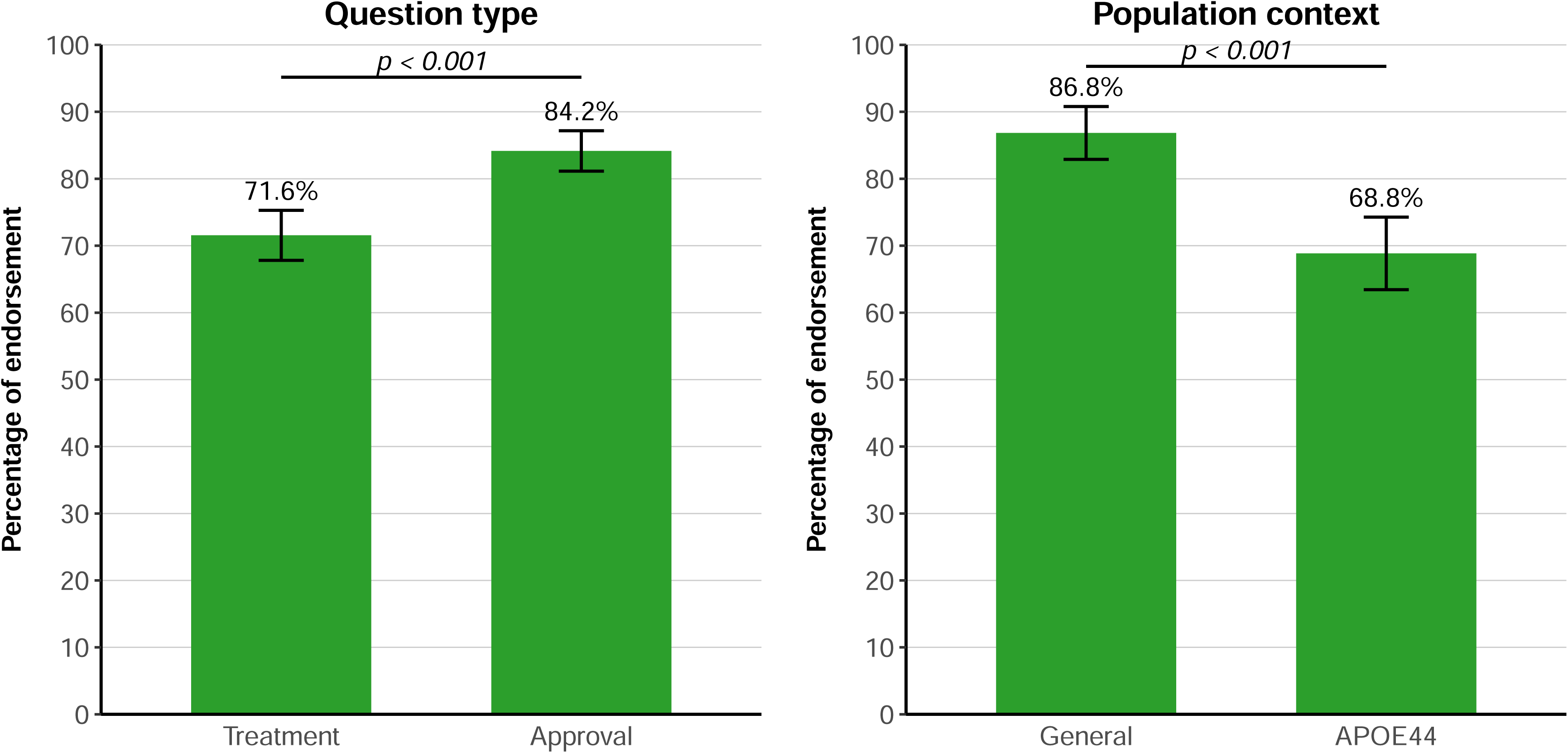
Endorsement by question type and population context. Endorsement rates were significantly higher for questions related to approval compared to those addressing treatment (left panel). Questions referring to the general early Alzheimer’s disease population received higher endorsement than those referring to APOE44 carriers (right panel). Error bars indicate 95% confidence intervals. Abbreviations: APOE44 = Homozygous carrier/carriers of the apolipoprotein 4 allele

### 3.6 Results by population context (general vs. APOE44)

Building on the observed pattern of higher endorsement in the general population context compared to the APOE44 carrier context, we categorized questions accordingly and carried out statistical analysis. Although endorsement rates were generally high, endorsement was statistically significantly higher for questions referring to the general AD population (approval: 92%; treatment: 82%) than for those referring specifically to APOE44 carriers (approval: 77%; treatment: 61%) (OR = 2.98, 95% CI [2.18, 4.09], p < 0.001) (Figure 4, right panel).

### 3.7 Survey findings across European regions (DNG, Austria, EADC)

When examining endorsement rates by population, the survey results were similar across regions. A statistically significant difference was found for the APOE44-specific approval question (p = 0.007). Post hoc pairwise comparisons (Holm-adjusted) revealed that this effect was primarily driven by a difference between Austria and EADC (p = 0.014), while other comparisons (DNG vs. Austria, DNG vs. EADC) were not significant after correction (p-values > 0.21). No significant differences between populations were observed for the other three questions (p-values > 0.17) (Table, Figure 5).

**Figure 5:**
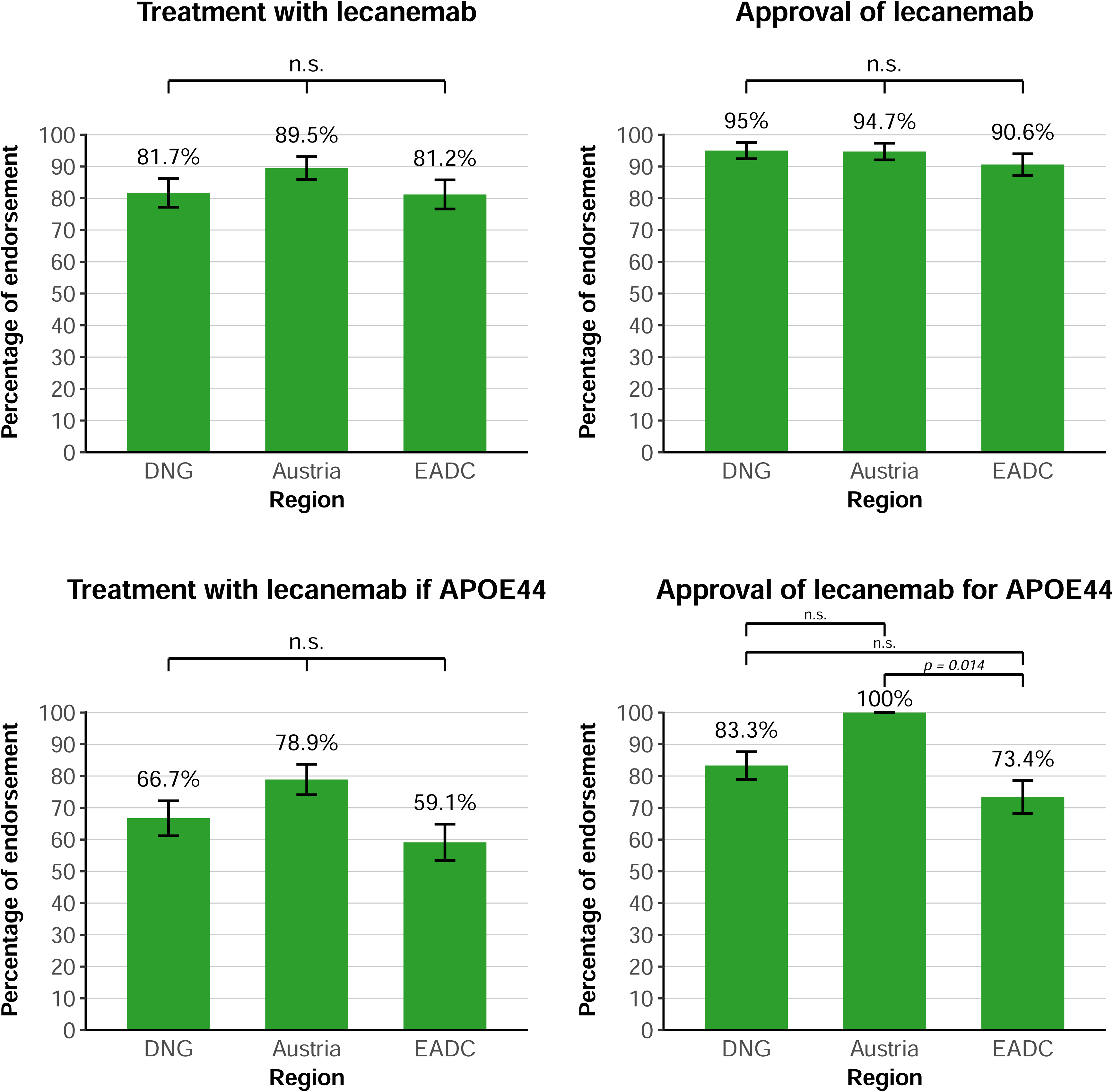
Similar endorsement rates across European regions. Endorsement was consistently high and did not significantly differ across regions for three of the four survey questions. For the question addressing approval for APOE44 carriers, a statistically significant difference was observed between Austria and EADC, potentially driven by complete response separation and the smaller sample size in the Austria population. Error bars indicate 95% confidence intervals. Abbreviations: APOE44 = Homozygous carrier/carriers of the apolipoprotein 4 allele; DNG = Deutsches Netzwerk Gedächtnisambulanzen (German Network of Memory Clinics); EADC = European Alzheimer’s Disease Consortium; n.s. = not statistically significant

### 3.8 Responses by timepoint (before vs. after CHMP’s positive opinion)

When stratified by timepoint (before vs. after the CHMP’s positive opinion), for the APOE44-specific approval question, endorsement was significantly lower after CHMP’s opinion (that excluded APOE44-carriers from approval) than before (before: 86.5%; after: 72.9%) (p = 0.025, OR = 0.42, 95% CI [0.18, 0.90]). No statistically significant differences were observed for the remaining three questions (treatment before vs. after: 86.5% vs. 80.2%; approval: 94.6% vs. 90.8%; treatment if APOE44: 68.9% vs. 58.5%; all p-values > 0.29) (Figure 6).

**Figure 6:**
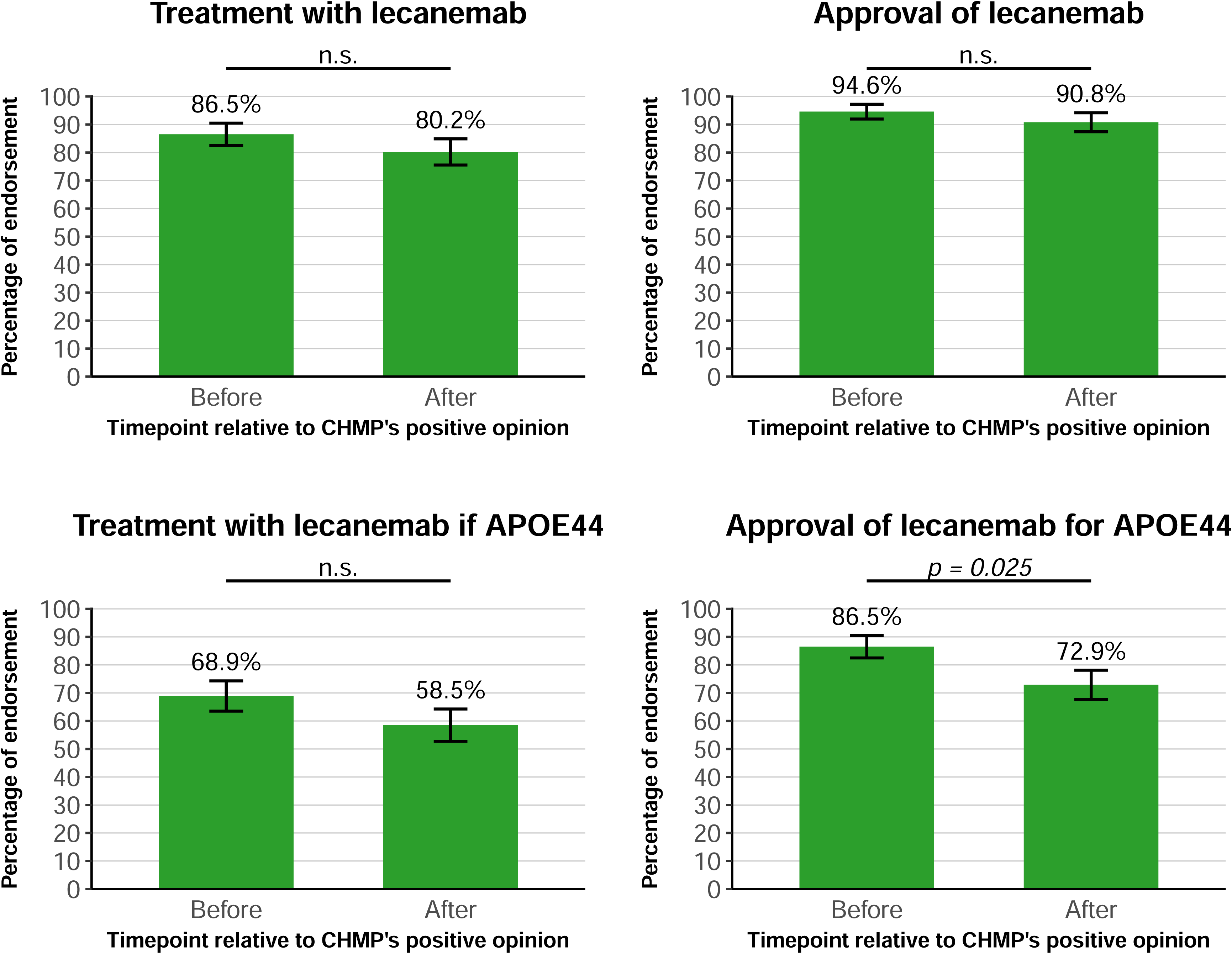
Endorsement relative to CHMP’s positive opinion regarding lecanemab approval in the European Union. For the approval question specific to APOE44 carriers (bottom right panel), endorsement was significantly lower after CHMP’s opinion (which excluded APOE44 carriers from the proposed label) than before. No significant differences were observed for the remaining three questions. Error bars indicate 95% confidence intervals. Abbreviations: APOE44 = Homozygous carrier/carriers of the apolipoprotein 4 allele; CHMP = Committee for Medicinal Products for Human Use; n.s. = not statistically significant

## 4 Discussion

In this European survey of patients with early symptomatic AD, we found strong overall support for lecanemab as both a treatment option and as a drug candidate for regulatory approval. Endorsement rates were significantly higher than expected by chance for all four survey questions. Notably, more than 90% of participants supported lecanemab’s approval in the EU, and over 80% indicated a personal wish to receive treatment with the drug. These findings provide important insights into patient preferences during a period when regulatory decisions were still pending in the EU [11]. The strong patient support for lecanemab observed in this survey stands in contrast to earlier regulatory hesitancy in the EU and aligns with the widespread reactions from scientific, medical and patient communities during the regulatory decision-making process, many of which expressed incomprehension at the initial negative CHMP opinion [13–21].

In addition to the consistently high endorsement for lecanemab use in the EU, our analysis revealed two robust response dynamics. First, approval-related questions consistently received higher endorsement than treatment-related questions. This pattern may reflect a distinction in how patients perceive personal treatment decisions versus broader regulatory policy. While some individuals may hesitate to pursue treatment for themselves, due to safety concerns, uncertainty about benefits, or personal values, they may nonetheless support access to lecanemab for others, emphasizing the importance of well-informed individual autonomy in treatment decisions [28]. This interpretation aligns with the ethical emphasis on patient-centered care and shared decision-making in AD [29,30], particularly as new therapies with complex risk-benefit profiles become available.

The second response dynamic was that questions framed in the context of the general AD population received higher endorsement than those referring specifically to APOE4 homozygotes. While the general concept of treatment or approval for early AD was met with broad support, the introduction of APOE4 homozygosity, an established risk factor for ARIA, appears to temper endorsement. Importantly, although endorsement was lower for APOE44-specific questions, the majority of participants still supported treatment and approval in this subgroup. This stands in contrast with the exclusion of APOE44 homozygotes from treatment [9,12] and indicates that patients may be willing to accept higher risks under certain conditions. This is in accordance with previous research showing that individuals affected by or at risk for AD may tolerate risks for slowing disease progression or maintaining autonomy for longer [31].

Notably, endorsement of approval for APOE44 homozygotes was not static over time. When stratified by timepoint, support for approval in this subgroup was significantly lower after the CHMP issued its opinion recommending approval for lecanemab while excluding APOE44 carriers from the proposed treatment indication. This decline, from 87% to 73%, suggests that regulatory decisions may shape public and patient perceptions, potentially reinforcing or amplifying concerns about treatment risks in this genetic subgroup. While the survey did not assess causal mechanisms, the observed temporal drop in endorsement may reflect the impact of regulatory communication on attitudes toward treatment eligibility and risk.

Overall, survey responses were broadly consistent across the participating European regions, with high levels of endorsement for lecanemab use observed in all groups. A statistically significant difference emerged only for the APOE44-specific approval question, where endorsement was higher in the Austrian subgroup compared to the EADC. This effect appears to have been driven by complete endorsement among Austrian respondents, combined with the smaller sample size of the Austria group, which may have amplified statistical contrasts. Apart from the observed difference between Austria and the EADC, no other group differences were found for the APOE44-specific approval question. Likewise, no significant regional differences emerged for the other three survey questions, supporting the overall consistency of patient attitudes toward lecanemab across European regions.

This study has limitations. The anonymous survey design ensured privacy but limited subgroup analyses, and the response rate could not be determined. Voluntary participation and physician-led recruitement may have introduced selection bias. Responses were limited to academic memory clinics and may not reflect the broader AD population. A known challenge in cognitively impaired populations with potentially reduced capacity of discernment is conveying information accurately. The survey’s brevity supported accessibility and focus on benefit–risk evaluation, but did not capture other potentially relevant aspects such as administration mode or frequency. Simplified, lay-friendly language to support accessibility may have resulted in wording that was not fully precise and led to varied interpretation. While the German and English versions were co-developed, other language versions relied on unstandardized AI-based translations. However, similar response patterns across populations may suggest that comprehension was consistent. Strengths of the survey include the high number of respondents, which increases the reliability and robustness of the findings. The inclusion of patients with biomarker-confirmed early symptomatic AD, rather than individuals who only express concerns about developing AD, ensures restriction of the survey to a population that is potentially eligible for lecanemab. The survey was conducted across multiple European memory clinics, allowing for a broader representation of attitudes beyond a single country or healthcare system.

The survey findings raise broader questions about the role of patients in regulatory decision-making processes. Beyond ethical considerations of autonomy and shared decision-making, the involvement of patients in the assessment of therapeutic value is increasingly recognized as a legal and institutional imperative [32]. The responses to the CHMP’s initial negative opinion by Alzheimer advocacy organizations and civil society stakeholders [19–21] underscore the relevance of including lived experience in regulatory deliberation. This is also consistent with principles of citizen science, which advocate for inclusive, transparent, and participatory approaches in areas of high public health relevance [33,34]. As disease-modifying therapies for AD continue to emerge [35], greater integration of patient perspectives may enhance the legitimacy, responsiveness, and acceptance of regulatory decisions.

An important consideration is the potential generalizability of these findings to other anti-amyloid therapies. As there is currently no robust evidence of differences in efficacy or safety among the internationally approved monoclonal amyloid-β antibodies lecanemab and donanemab, the preferences expressed in this survey may therefore similarly apply to donanemab, which demonstrated efficacy in a positive phase 3 trial [36] and has subsequently been approved in multiple regions worldwide, including the USA [37], Japan [38], China [39], and the UK [40]. On March 27, 2025, the CHMP issued a recommendation to refuse approval of donanemab in the EU [41].

In summary, this survey provides timely and unique insights into the attitudes of patients with early symptomatic AD in Europe toward lecanemab and anti-amyloid therapies more broadly. The consistently high levels of support observed, both for treatment and for regulatory approval, highlight that patients value the availability of therapeutic options, even when associated with complex risk–benefit profiles. The results further suggest that patients are willing to engage with nuanced treatment decisions, including those involving genetic risk, and that they may accept elevated risks under specific conditions. As regulatory agencies continue to evaluate emerging therapies for AD, these findings underscore the importance of integrating patient perspectives into benefit–risk assessments and policy decisions. Looking ahead, future research should explore how preferences evolve with real-world treatment experience, and how best to operationalize meaningful patient involvement in regulatory, clinical, and ethical frameworks surrounding disease-modifying therapies in Alzheimer’s disease.

## Data Availability

All data produced in the present study are available upon reasonable request to the authors.

## Acknowledgements

We thank the study participants for their effort to take part in the survey. We thank the physicians of the EADC memory clinics, the DNG and of contributing memory clinics in Austria who contributed by identification of participants fulfilling the eligibility criteria for this investigation.

## Consent statement

Consent was not required in this anonymous survey.

## References

[1] Van Dyck CH, Swanson CJ, Aisen P, et al. Lecanemab in Early Alzheimer’s Disease. N Engl J Med. 2023;388(1):9–21.

[2] https://www.fda.gov/news-events/press-announcements/fda-converts-novel-alzheimers-disease-treatment-traditional-approval; accessed May 12, 2025

[3] https://www.eisai.com/news/2023/news202359.html; accessed May 12, 2025

[4] https://www.eisai.com/news/2024/news202403.html; accessed May 12, 2025

[5] https://www.eisai.com/news/2024/news202436.html; accessed May 12, 2025

[6] https://www.eisai.com/news/2024/news202449.html; accessed May 12, 2025

[7] https://www.eisai.com/news/2024/news202451.html; accessed May 12, 2025

[8] https://www.eisai.com/news/2024/news202460.html; accessed May 12, 2025

[9] https://www.gov.uk/government/news/lecanemab-licensed-for-adult-patients-in-the-early-stages-of-alzheimers-disease; accessed May 12, 2025

[10] Honig LS, Sabbagh MN, van Dyck CH, et al. Updated safety results from phase 3 lecanemab study in early Alzheimer’s disease. Alzheimers Res Ther. 2024;16(1):105.

[11] https://www.ema.europa.eu/en/medicines/human/EPAR/leqembi; accessed May 12, 2025

[12] https://ec.europa.eu/newsroom/sante/items/879055/en

[13] Jessen F, Kramberger MG, Angioni D, et al. Progress in the Treatment of Alzheimer’s Disease Is Needed - Position Statement of European Alzheimer’s Disease Consortium (EADC) Investigators. J Prev Alzheimers Dis. 2024;11(5):1212–1218.

[14] https://www.sciencemediacentre.org/expert-reaction-to-ema-decision-on-lecanemab-for-alzheimers-disease/

[15] The Lancet. Divisions over lecanemab: keeping an open mind. Lancet. 2024 Sep 21;404(10458):1077

[16] https://dgn.org/artikel/dgn-bedauert-chmp-empfehlung-gegen-die-ema-zulassung-des-ersten-alzheimer-antikorpers-in-europa; accessed May 12, 2025

[17] https://www.dgppn.de/presse/pressemitteilungen/pressemitteilungen-2023/alzheimer-behandlung-antikoerper-therapie-darf-betroffenen-nicht-verwehrt-werden.html; accessed May 12, 2025

[18] https://www.dzne.de/en/im-fokus/meldungen/2024/the-decision-is-incomprehensible-statements-of-dzne-experts-on-the-refusal-of-lecanemab-by-european-medicines-agency/; accessed May 12, 2025

[19] https://www.alzheimer-europe.org/news/26-july-alzheimer-europe-regrets-negative-european-medicines-agency-opinion-lecanemab-may?language_content_entity=en; accessed May 12, 2025

[20] https://www.alzheimer-europe.org/news/alzheimer-europe-responds-negative-opinion-lecanemab-european-medicines-agency; accessed May 12, 2025

[21] https://www.alzheimer-europe.org/news/our-member-organisations-react-ema-opinion-lecanemab; accessed May 12, 2025

[22] https://www.eisai.com/news/2024/news202455.html; accessed May 12, 2025

[23] https://www.zeit.de/gesundheit/2024-07/alzheimer-therapie-lecanemab-patienten-studie; accessed May 12, 2025

[24] https://english.elpais.com/science-tech/2024-09-25/a-scientific-rebellion-to-get-europe-to-approve-a-controversial-alzheimers-drug.html; accessed May 12, 2025

[25] https://www.ema.europa.eu/en/medicines/human/EPAR/leqembi; accessed May 12, 2025

[26] https://deutschesnetzwerkgedachtnisambulanzen.clubdesk.com/verein/sektionen; accessed May 12, 2025

[27] https://eadc.online/member-list/#1669493220960-e5696535-de6f; accessed May 12, 2025

[28] Olchowska-Kotala A, Strządała A, Barański J. Patients’ Values and Desire for Autonomy: An Empirical Study from Poland. J Bioeth Inq. 2023 Sep;20(3):409–419.

[29] Daly RL, Bunn F, Goodman C. Shared decision-making for people living with dementia in extended care settings: a systematic review. BMJ Open. 2018 Jun 9;8(6):e018977.

[30] Miller LM, Whitlatch CJ, Lyons KS. Shared decision-making in dementia: A review of patient and family carer involvement. Dementia (London). 2016 Sep;15(5):1141–57.

[31] Roldan Munoz S, de Vries ST, Lankester G, et al. Preferences about Future Alzheimer’s Disease Treatments Elicited through an Online Survey Using the Threshold Technique. J Prev Alzheimers Dis. 2023;10(4):756–764.

[32] https://www.ema.europa.eu/en/partners-networks/patients-consumers; accessed May 12, 2025

[33] Groot B, Abma T. Ethics framework for citizen science and public and patient participation in research. BMC Med Ethics. 2022; 13;23(1):23.

[34] Robinson L.D., Cawthray, J.L., West, S.E., et al. Ten principles of citizen science. In S. Hecker, M. Haklay, A. Bowser, Z. Makuch, J. Vogel, & A. Bonn. Citizen Science: Innovation in Open Science, Society and Policy. London, UCL Press. 1–23.

[35] Cummings J, Zhou Y, Lee G, et al. Alzheimer’s disease drug development pipeline: 2024. Alzheimers Dement (N Y). 2024 Apr 24;10(2):e12465.

[36] Sims JR, Zimmer JA, Evans CD, et al. Donanemab in Early Symptomatic Alzheimer Disease: The TRAILBLAZER-ALZ 2 Randomized Clinical Trial. JAMA. 2023 Aug 8;330(6):512–527.

[37] https://www.fda.gov/drugs/news-events-human-drugs/fda-approves-treatment-adults-alzheimers-disease; accessed May 12, 2025

[38] https://investor.lilly.com/news-releases/news-release-details/lillys-kisunlatm-donanemab-azbt-approved-japan-treatment-early; accessed May 12, 2025

[39] https://investor.lilly.com/news-releases/news-release-details/lillys-kisunlatm-donanemab-azbt-approved-china-treatment-early; accessed May 12, 2025

[40] https://www.gov.uk/government/news/donanemab-licensed-for-early-stages-of-alzheimers-disease-in-adult-patients-who-have-one-or-no-copies-of-apolipoprotein-e4-gene; accessed May 12, 2025

[41] https://www.ema.europa.eu/en/medicines/human/EPAR/kisunla; accessed May 12, 2025

